# A unified activities-based approach to the modelling of viral epidemics and COVID-19 as an illustrative example

**DOI:** 10.1101/2020.06.10.20127597

**Authors:** Yulii D. Shikhmurzaev, Vladislav D. Shikhmurzaev

**Affiliations:** School of Mathematics, University of Birmingham, Birmingham B15 2TT, United Kingdom; Clinical City Hospital #52, 3 Pekhotnaya Str., Moscow 123182, Russia

**Keywords:** Coronavirus, Social map and restrictions, Transmission pattern

## Abstract

A new approach to formulating mathematical models of increasing complexity to describe the dynamics of viral epidemics is proposed. The approach utilizes a map of social interactions characterizing the population and its activities and, unifying the compartmental and the stochastic viewpoints, offers a framework for incorporating both the patterns of behaviour studied by sociological surveys and the clinical picture of a particular infection, both for the virus itself and the complications it causes. The approach is illustrated by taking a simple mathematical model developed in its framework and applying it to the ongoing pandemic of SARS-CoV-2 (COVID-19), with the UK as a representative country, to assess the impact of the measures of social distancing imposed to control its course.

## 1 Introduction

The key difference between an animal and a human population relevant to the modelling of epidemics of communicable diseases is that the latter lives in an artificial environment sustained by its own effort. This environment, even with measures of social distancing in place, still requires a certain level of interpersonal communication structured in a way specified by economics and the corresponding differentiation of social roles. Consequently, policy recommendations following from a model of an animal-like population, where self-isolation for as long as it takes invariably stops any epidemic at any stage, become inapplicable given that self-isolation — even for a few days — executed just by those maintaining the food supply chain would lead to a total collapse of the living environment and disintegration of society. Thus, for a mathematical model of the epidemic dynamics to be useable in practice, it should differentiate individuals first of all with regard to their socioeconomic roles and account for the interpersonal contacts associated with them.

A review of currently used approaches to the modelling of epidemics in human populations, e.g., [1], shows that they stem from the works treating the population as composed of identical interchangeable individuals with no socioeconomic functions, i.e. essentially animals. Indeed, the compartmental models originating from the seminal papers of Kermack and McKendrick [2–4] break the population into groups (‘compartments’) differentiated only with respect to the disease (Susceptible, Infective, Recovered, abbreviated to SIR, or, when Exposed are added, to SEIR). In these models, often referred to as ‘standard’, the individuals forming the population have no other social roles apart from transmitting the infection and recovering or dying from it.

Stochastic models, built on the mathematics of the Galton-Watson process [5–8], follow [9] and the developments summarized in [10] with the transmission dynamics considered as a graph of interactions with probabilities of transmission ascribed to the edges. More realistic with regard to the probabilistic nature of the disease transmission, this approach again makes no distinction between human and animal populations nor does it describe the population as a whole.

The assumption that the population is homogeneous and socially structureless has been enshrined [11] in the notion of the basic reproduction number *R*_0_ defined as ‘the expected number of disease cases produced by a typical infected individual in a wholly susceptible population over the full course of the infectious period’ [1]. This notion as well as the current reproduction number *R* used to characterize the epidemic in its course are helpful statistical indicators but, should they be tested literally in a real-life (albeit thought) experiment, they would spectacularly vary depending on what socioeconomic group this ‘typical individual’ belongs to.

The limitations of the homogeneous population assumption have long been recognized and a number of works [12–16] aimed to address them were published, gradually evolving from a somewhat ad-hoc idea of ‘superspreaders’ [17, 18] to a generic approach where the population is divided into an arbitrary number of groups, each with the SEIR structure, interacting in an arbitrary number of locations (‘patches’) [19], with the times spent by member of each group in each location characterized by a set of mixing matrices. The latter somewhat abstract approach is an extension on earlier works in this direction [20–23] where particular, more disease-specific, situation were considered. A shortcoming of the developed approach, apparent once it is reduced to its simplest variant, is that, according to it, a representative susceptible comes in contact only with *one* person in a given ‘patch’, so that the probability of coming across an infective is the ratio of the number of infectives to the total population in this patch. Neither the groups nor the patches are linked with social activities and this makes it difficult to specify the matrices involved for a practical application of the model, and the basic SEIR scheme makes it problematic to account for the infectiveness ‘profile’ of a disease in question. The latter aspect is of particular importance in the situations where many cases go asymptomatically with varying infectivity and/or if the infection has a long latent period.

An essential element in the overall dynamics of an epidemic is the recovery process which, in the case of a viral infection, brings back into the community the individuals who are now immune to the disease and hence contributes towards building ‘herd immunity’. In its turn, the recovery process is inseparable from the functioning of the healthcare system so that the epidemics-related role of the healthcare system also has to be modelled mathematically, at least on a basic level. In short, a realistic mathematical model intended to describe epidemics of communicable diseases requires a systematic approach able to incorporate (i) the specifically human form of organization of society and the social groups determined by it, (ii) the ‘epidemiological profile’ of the disease accounting for both the time scales characterizing its stages and the associated variations in the probabilities involved in the spreading and recovery process, and (iii) the functioning of the health care system in terms of effectiveness and availability of medical help.

In the present work, we develop a systematic approach to constructing mathematical models of increasing complexity based on the map of social interactions characterizing the population. The approach brings together the compartmental and stochastic aspects of the epidemic dynamics and offers a natural framework for directly incorporating the epidemiologically relevant features of the disease, both associated with the virus and the complications it may cause, as well as the role played by the healthcare system. The approach is illustrated by considering the simplest mathematical model formulated in its framework. The model is described in the form of an algorithm which provides a practical user-friendly instrument for the description of the course of epidemics and, in particular, allows one to model the impact of restrictions and social distancing measures that could be imposed to curb an outbreak. The algorithm is applied to the ongoing epidemic of COVID-19 as a convenient illustrative example.

The paper is organized as follows. In Section 2 we outline the reasons behind the choice of the mathematics to be used and the caveats regarding the use of observational data. In Section 3, the conceptual framework for the modelling is presented, and in Section 4 we describe an algorithm of how a day-by-day evolution of the characteristic parameters of the epidemic can be computed. In Section 5, the algorithm is applied to the ongoing epidemic of COVID-19 in the UK, where we use the observational data to analyze the impact of social distancing both retrospectively and for the future. Finally, in Section 6, some directions of generalization of the developed approach are discussed.

## 2 Preliminary remarks

### 2.1 Continuous time versus discrete time

The starting point in developing a mathematical model in any field is the choice of mathematics to be used. This, generally nontrivial, question is usually resolved either traditionally, following successes of modelling in the field, or by default, bringing in and adapting the mathematics one is familiar with. However, it is the choice of the appropriate mathematics that largely determines the outcome of the modelling as the concepts needed for the description have to be cast in the form required by the chosen mathematics.

In the epidemiology of the compartmental approach, following Kermack and McKendrick [2–4], the outcome of the modelling is usually expected to take the form of (ordinary) differential or integro-differential equations for the evolution in time of the dependent variables characterizing the number of people in different compartments. This expectation involves an implicit assumption that the time is a continuous independent variable and the dependent variables are differentiable functions of it. What this assumption means is that the time scale of ‘elementary processes’ and the time scale on which the system as a whole evolves are well separated so that one can introduce an intermediate time scale of averaging, large compared with the former and small compared with the latter, for the correspondingly averaged quantities to make sense. A well known example of this type of modelling can be found in continuum mechanics [24] where one deals with differential equations, both in the spatial variables and time, as the length and time scales associated with molecular interactions (‘elementary processes’) are negligible compared with the length and time scales on which averaged quantities characterizing the medium evolve.

This is manifestly not the case with the dynamics of epidemics where the ‘elementary processes’ of susceptibles turning into infectives occur on the same time scale as the time scale on which the numbers characterizing the epidemic macroscopically evolve: these numbers simply accumulate individual acts of contracting the disease. To put it simply, the epidemiological status of an individual, if changes, does so on a daily basis as well, and this is also the time scale on which the variables characterizing the epidemic as a whole evolve. This means that for the epidemic the time should not be treated as a continuous variable; it is discrete, measured in days and should be treated as such in the model. Hence, for the epidemic dynamics one should choose the mathematics of the difference, not differential, equations, with the time step of 1 (day). Importantly, the epidemic’s dynamics is monitored on a daily basis and the clinical status of those affected is also reported daily in the epidemics statistics so that the mathematics of the difference equations provides a natural framework for both inputting the medical data and comparing the model with observations.

By contrast, the virus replicates on a time scale much shorter than 24 hours, and therefore, in order to describe how it changes the daily infectivity profile of the infection carrier, one has to use the continuous time-based description and then discretize it as required.

### 2.2 Uncertainty in observational data

The main characteristic of an epidemic to be described by a mathematical model is the number of those who contracted the disease and how this number evolves with time. For the model to be verified by comparing the results it predicts with observational data, ideally, (a) the whole population should be tested on a daily basis, (b) those tested positive but showing no symptoms should not be informed for them not to change their social behaviour and continuing to spread the disease as if not tested, and (c) the social behaviour of the population should not be influenced by the information campaign about the ongoing epidemic broadcast by the media. Obviously, for a serious epidemic, like the ongoing COVID-19 pandemic, none of these conditions, ideal for the epidemics modeller, can be satisfied. The cases reported in the epidemics statistics refer not to those infected; the data refer to those *diagnosed*. The number of diagnosed is a mix of those tested positive who showed clinical symptoms and sought medical help and those of the asymptomatic infectives who happened to be covered by the testing programme and tested positive. The former had already withdrawn from daily activities because of being ill and hence no longer spread the disease whilst the latter have to withdraw from the daily routine and self-isolate because of the result of testing. In other words, the reported epidemiological data are, like in quantum mechanics, affected by the way they are collected. The wider the testing programme, the more asymptomatic infectives it turns into those with a diagnosis and hence the more it makes the collected data a mix and the subsequent course of the epidemic influenced by it as a factor external to the epidemic as such. A related issue is reliability of the tests themselves. In the case of a new virus, the testing methods evolve as the epidemic unfolds which often makes earlier data underpredictive. In short, from the modelling viewpoint, the reported epidemiological data should be taken with caution, and one should begin with clarifying what the reported cases actually represent.

All this is illustrated in Fig. 1, where we plot, on the linear and a semilogarithmic axes (left and right, respectively), the dynamics of diagnosed cases of COVID-19 in the UK, Germany, Japan and Russia. The initial growth of the number of cases in an epidemic is expected to be exponential, i.e. linear on the semilogarithmic axes, with the slope determined by the virulence of the virus and the communication pattern of the population. As can be seen, the data for the UK, Germany and Russia show the expected exponential trend (Fig. 1, right) with the same slopes on the semilogarithmic axes. However, the data for Japan, while also showing the exponential trend, has a markedly different slope indicating some systemic reason underneath.

**Fig. 1.**
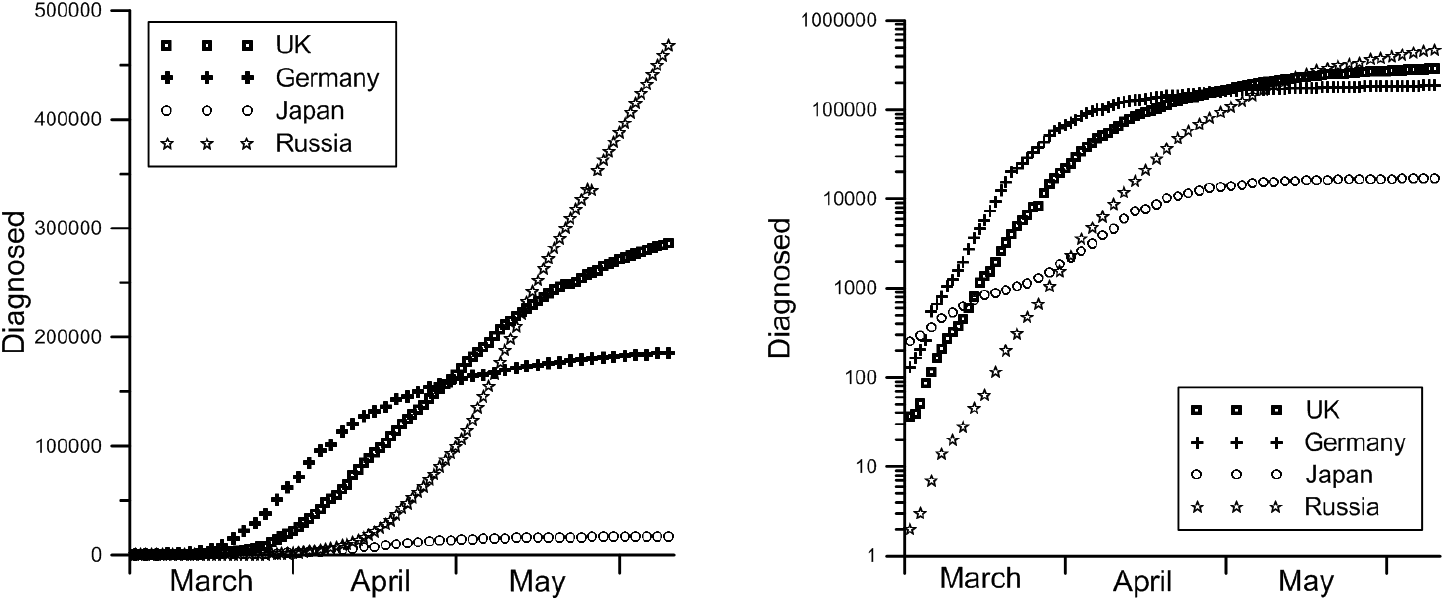
Diagnosed cases of COVID-19 in 4 countries in the spring of 2020. Left: linear plot; right: semi-logarithmic plot.

An important point to be noted here is that the data for the UK and Russia, albeit looking similar, represent essentially different cases. In the UK, the guidance issued to the population was (and, for the time being, is) that those who experience clinical symptoms of COVID-19 (fever, dry cough, etc) should self-isolate for seven days, get advice online, see what comes and call for medical help if matters get worse. Thus, the UK data gives a relatively clean sample, with the reported cases representing essentially those with complications of the infection, whilst the set of those with no or mild to moderate symptoms can be much wider. By contrast, in Russia the guidance is to call the ambulance at the first symptoms or even suspicion and, in addition, a very wide testing programme, with blanket coverage of large segments of the population, is in place, with hospitals financially incentivized to bring asymptomatic infectives into the fold. As a result, the Russian data is of much less value for the modeller as it strongly depends on the testing programme as an external factor, with the number of cases increasing as the testing programme widens.

The situation with mortality, which also could be used to test the model, is much the same. Although death is an undeniably certain outcome, the reported numbers comprise those who died, generally speaking, *from* the virus and those who died *with* the virus. In other words, they put together those who died from complications directly caused by the virus as well as those who had the virus but died from conditions, possibly exacerbated by the infection, but not directly caused by it. Separation of mortality from ‘co-mortality’ reduces the uncertainty but only to a certain degree. The mortality rate is implicitly affected by the testing programme as well since early diagnostics generally reduces mortality. In any case, the reported mortality data should not be considered from the viewpoint of natural mortality from the disease since the data, especially in the late stages of an epidemic, strongly depend on the efficiency of the healthcare system and evolve as more experience is gained. From the modelling viewpoint, the probability of dying from complications caused by the virus without medical help, needed in a closed model in one form or another as a baseline from which the efficiency of medical treatment is to be measured, is obviously never available.

The above issues with the mortality data are illustrated in Fig. 2 showing mortality from COVID-19 for the same four countries as in Fig. 1. The reported cases are shown both as the raw data (left) and as a dead-to-diagnosed ratio (right). It is noteworthy that the relatively low mortality rate recorded in Russia is consistent with the 2.84% mortality rate reported for Wuhan, China in an early study [25], whilst a high mortality rate reported in the UK, even compared with the world average (currently below 7%) and outside the 4–11% range for hospitalized adult patients elsewhere [26], is yet to be explained.

**Fig. 2.**
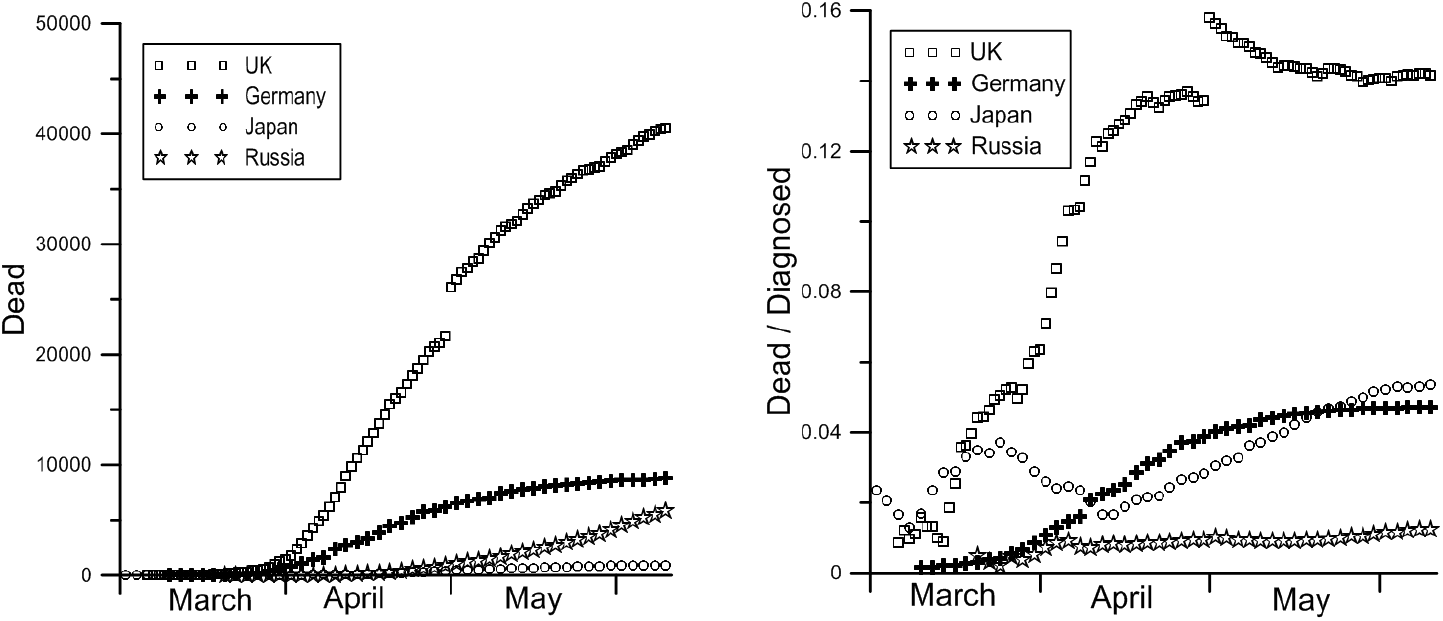
Deaths from COVID-19 in 4 countries in the spring of 2020. Left: absolute numbers; right: the dead-to-diagnosed ratio.

Thus, the reported epidemiological data bears a considerable degree of uncertainty and as a basis for quantitative testing of a mathematical model has a somewhat limited value. An informative indicator is the slope of the initial linear part in the semilogarithmic plot of the diagnosed-versus-time curve (Fig. 1, right) reflecting infectivity of the virus. It is this slope that can be interpreted in terms of *R*_0_. The main qualitative feature that a realistic model should be able to capture is the effect of the measures of social distancing imposed on the population and the associated time scales.

## 3 Conceptual framework

### Generic clinical picture

Consider an epidemic caused by a viral infection. Schematically, the picture we will have in mind is as follows. After contracting the virus, an individual becomes a carrier (‘infective’) and, for the duration of the latent (incubation) period characteristic of the virus but with individual variations within a certain range, does not feel the infection. Then, the dis-ease either continues asymptomatically, with the carrier remaining infective to others during this time, and eventually clears off or, alternatively, after the incubation period, the carrier falls ill showing some or all clinical symptoms of the disease. In the latter case, the individual in question either recovers on his/her own in due course, at home or possibly spending some time in hospital, or develops complications. In the latter case, in the terminology we will use here, he/she becomes hospitalized/critically ill and either recovers, this time on a time scale associated with the complications and generally different from that characterizing the viral infection, or dies. In the model, we quantify this branching scenario and introduce the probabilities that control it.

In what follows, we discretize the time into days, as opposed to using the continuous time adopted in the dynamical system-type modelling, where meaningful events, like transmission of the disease, happen continuously day and night. As we will see, the natural time scale we use here makes transparent both the meaning of the model’s parameters and comparison of the results with the real-life data; if necessary, the model can be adapted for a shorter time step although such a move is nontrivial. In dealing with the relevant functional dependencies, we will represent them in a simple functional form intended to capture the trends and elucidate the influence of parameters; where the relevant data become available with higher accuracy than the overall accuracy of the model, these dependencies can be replaced with the empirical data without any changes to the approach itself. As tests have shown, the model is rather robust with respect to the exact shape of the functions used; what matters is the parameters determining the scales.

### Structure of community

The first step in our approach is to break the population into categories relevant to (a) the functioning of the community (city, country) and (b) the infection in question in terms the category’s communication pattern and the role in combatting the disease. In the simplest model used here to illustrate the approach, we have the following five categories:

*W* - ‘workers’, i.e. those whose work involves interaction primarily/exclusively within their own professional community.

*S* - service providers whose work involves communication with people from different walks of life, i.e., in terms of the present model, with other categories, as well as with fellow service providers.

*M* - medical personnel involved in combatting the epidemic in question and the associated support staff providing infrastructure and logistics.

*R* - retired/elderly.

*U* - unemployed and other economically inactive people.

The list of categories can be expanded, for example, by splitting *W* and *S* into groups such ‘key workers’ whose work is vital for maintaining the community during an epidemic and ‘non-essential workers’ whose work could be suspended or performed remotely. Further generalization could be by splitting each category into gender and age groups, differentiating the population by socializing patterns, vulnerability to the infection and location, aggregating them into different types of households, etc.

It should be emphasized that the five-category social structure used here differs from the traditional splitting into ‘manufacturing’ and ‘service’ industries adopted in statistical datasets reported, for example, by the Office for National Statistics (UK). Traditionally, ‘service industries’ include the retail sector, financial sector, public sector business administration, leisure and cultural activities. However, viewed through the prism of epidemiology and the above categories, a substantial part of people involved in traditional ‘service industries’ belong to our category *W*, not *S*. For example, the education sector comprises not only those who provide education but also those receiving it and therefore should be attributed to *W* as interacting within their own professional community. Furthermore, in the service sector as such a substantial part of jobs are back office jobs not dealing with customers and hence again belonging to *W*. The frontline medical personnel are singled out into a separate category (*M*) whilst others are split between *W* (lab and admin staff) and *S* (medics not dealing with the infection in question). The figures in categories *R* and *U* can be taken directly from statistical reports.

On day *n* of the epidemic, the people belonging to category *C* (*C* = *W, S, M, R, U*) can be in different states with regard to the disease which we will label as follows:

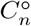 - ‘susceptible’, i.e. healthy but not immune to the virus.

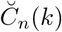 - ‘infected’, i.e. those who are carrying the virus for *k* days but showing no/negligible symptoms.

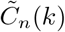 - ‘symptomatic’ for *k* days, no longer participating in social activities but with no complications.

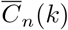 - ‘hospitalized’, i.e. those who for *k* days suffer from complications caused by the virus.

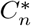 - ‘recovered’, i.e. healthy and now immune to the virus.

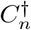 - dead.

The susceptible, infected and recovered people take part in their usual daily activities. The symptomatic ones feel unwell and no longer participate in outdoor activities but will recover from the virus in due course on their own unless the virus causes complications in which case they turn into ‘hospitalized’. The hospitalized need medical help and will either recover, albeit on a time scale different from that of recovery from the virus, or die.

Below, the letters *W, S, M, R* and *U* equipped with the signs corresponding to the state with regard to the infection will refer to the number of people in each category and state but, when used in the subscript (e.g. in *F*_(*a,C*)_ below), they will be simply indices corresponding to the number of the category in the list above (i.e. *W* = 1, *S* = 2, *M* = 3, *R* = 4, *U* =5). As we will see, this dual use of symbols makes the reading easier.

It should be noted that, in general, the number of people in the population as a whole is not fixed as there can be flows of people in and out, e.g. medical staff can be brought in from the outside or dispatched to help elsewhere, tourists can come back home, healthy or otherwise, there can be travel between communities if the model is applied, say, to a city or some part of the country, etc. Such flows can be easily accounted for, and in what follows, for simplicity, we neglect them.

### Social activity

The second step in our approach is to identify the activities where communication between individuals takes place on a daily basis. Here the term ‘daily basis’ is understood as an average over a characteristic interval, e.g. a week. For the simplest illustrative model, we introduce the following four main activities:

*t* - commuting by public transport;

*w* - work, including attending schools and universities;

*e -* all interactions with service providers, including when taking care of the essentials (food, pharmacies, etc);

*s* - socializing.

The list of activities could again be expanded alongside the list of categories. For example, ‘transport’ can be split according to the means of transport, ‘work’ can be split into sectors or even down to companies, ‘essentials’ can be split into groups according to location of the relevant facilities, and ‘socializing’ can be broken down into groups, e.g. corresponding to the categories of people involved.

### Map of social interactions

Given the social structure of the population and the list of activities its members are involved in, we need to characterize daily participation of each category of people in each of the activities with regard to (a) what fraction of the people in this category simultaneously takes part in the activity and (b) how many interactions on average and with whom they have during this activity.

In order to characterize these factors, we introduce the following:

*F*_(*a,C*) -_ the fraction of the population from category *C* (*C* = *W*, …, *U*) simultaneously taking part on a daily bases (on average) in activity a, (*a* = *t*, …, *s*).

*K*_*t*_ *-* the number of daily contacts in public transport.

*K*_*C*_ - the number of daily contacts with colleagues for *C* = *W* or with infection carriers for *C* = *M*.

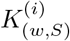 - the number of daily contacts service providers have with colleagues (*i* = 1) and with customers (*i* = 2).

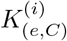 - the number of daily contacts people from category *C* (*C* = *W*, …, *U*) have as customers with other customers (*i* = 1) and with service providers (*i* = 2).

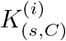 - the number of social daily contacts a typical individual from *C* (*C* = *W*, …, *U*) has within its own category (*i* = 1) and outside (*i* = 2).

It should be noted here that ‘contact’ here is defined with respect to the infection to be modelled as in the same situation socially this number will be different, for example, for influenza and for smallpox. The generalization here could go along the lines of the viral load but we will not discuss it here.

From an epidemiological viewpoint, the 4*×*5 matrix *F*_(*a,C*)_, with the entries varying between 0 and 1, together with the set 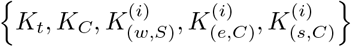, *C* = *W*, …, *U* ; *i* = 1, 2 characterizing the average number of contacts in different situations form the simplest ‘map of social interactions’ or a ‘social signature’ of the community with regard to a particular infection corresponding to the social structure and activities introduced above. Sociological surveys show that this ‘signature’, i.e. the pattern of social behavior, is remarkably stable, and, as we will see below, it is the elements of this ‘signature’ that become subject to restrictions imposed at the time of an epidemic. Strictly speaking, one needs separate maps of social interactions for the work days and weekends/holidays but here, for simplicity, we will not consider this generalization.

The matrix *F*_(*a,C*)_, which we will call ‘participation matrix’, should not be confused with ‘mixing matrices’ featuring in different forms in a number of models, e.g. [27–31]: in our approach, the first index refers to an activity and the second indicates a category of people whilst in mixing matrices both indices refer to the groups whose representatives come in contact.

### Infection

We will characterize the viral infection and the complications that may follow using the following parameters. First, we need to characterize the viral infection itself:

*t*_1_ - the average duration of the incubation/latent period of the viral infection, i.e. the period between contracting the virus on ‘day 0’ and its clinical symptoms if they are to follow.

σ_1_ - the standard deviation from *t*_1_ due to individual variability.

*n*_0_ - the average duration of the viral infection in an asymptomatic infective.

*n*_1_ - the recovery period for a symptomatic case, i.e. the period from the appearance of clinical symptoms to recovery.

*p*_0_ - the probability of contracting the virus from a virus carrier at the end of the latent/incubation period of the latter.

*p*_1_ - the probability of falling ill, i.e. showing clinical symptoms of the viral infection.

Then, we need to characterize the complications caused by the virus and the associated mortality:

*t*_2 -_ the average time from clinical symptoms of the viral infection to the onset of complications.

σ_2_ - the standard deviation from t2 due to individual variability.

*n*_2_ - the recovery period from complications.

*p*_2_ - the probability of developing complications, i.e., becoming critically ill.

*t*_*m*_ *-* the average time from the onset of complications to death for non-survivors.

σ_*m*_ *-* the standard deviation on *tm* due to individual variability.

*p*_*m*_ - the probability of dying in the absence of medial help (baseline probability).

The probabilities *p*_0_, *p*_1_, *p*_2_ and *p*_*m*_ introduced above provide the scales but to consider the epidemic dynamics we need to write them into probability distributions:.

### Probabilistic profile of the disease

As outlined above, there is a cascade of possible events corresponding to the course of the disease for an individual, and to describe it we introduce the following distributions.

*P*_0_(*k*) is the probability with which an infective who contracted the virus *k* days ago will transmit it to a susceptible at a socially close contact

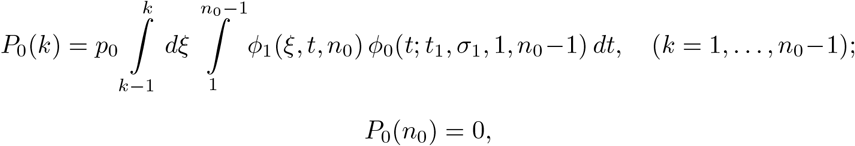

where

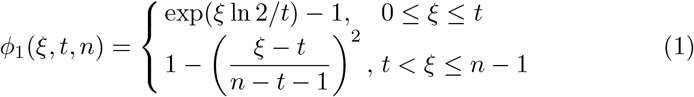

is the schematically represented virulence profile of the virus. The function *ϕ*_1_ is a typical profile accounting for an exponential growth during the incubation period and an algebraic decay following the immune response whilst the function *ϕ*_0_, taken in the form of a truncated normal distribution

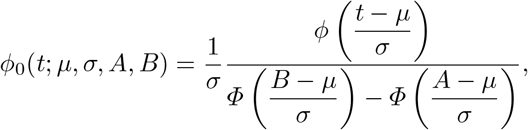

where

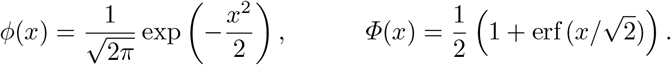

accounts for individual variability. The set of probabilities *P*_0_(*k*) implies that the infection carrier is not contagious on day 0 and day *n*_0_ of the viral cycle. Should the empirical data on *P*_0_(*k*) be available, it can be used instead of the dependence we use here.

*P*_1_(*k*) is the probability that an infective who contracted the virus *k* days ago falls ill, i.e. shows clinical symptoms, and hence withdraws from regular daily activities. For *P*_1_(*k*) we assume a discretized truncated normal distribution,

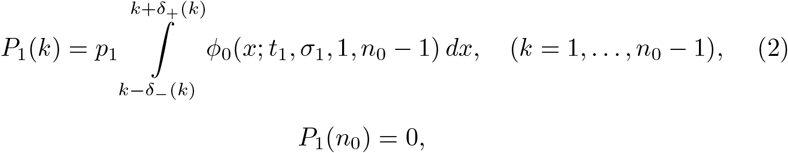

where *δ*−(1) = *δ*+(*n*_0_ − 1) = 0 and

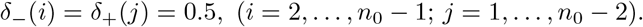

*P*_2_(*k*) is the probability that *k* days after showing clinical symptoms of the viral infection the person develops complications, i.e. becomes critically ill. This profile of the viral infection with respect to the complications it causes depends on the particular virus and can include several waves corresponding to different complications. Here, we will use a simple one-wave pattern described by the discretized truncated normal distribution similar to that in (2):

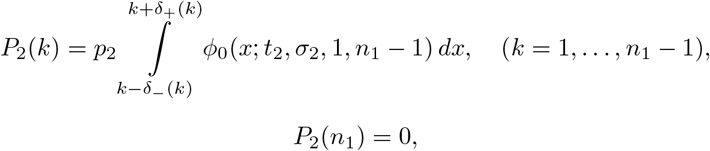

where again *δ*_−_(1) = *δ*_+_(*n*_1_ − 1) = 0 and

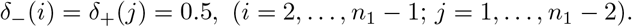

*P*_3_(*k*) is the probability of dying after being critically ill for *k* days. For this probability we will again use the discretized truncated normal distribution:

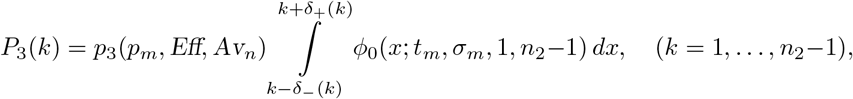

where *δ*−(1) = *δ*+(*n*_2_ − 1) = 0,

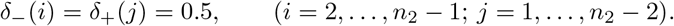

The probability *p*_3_(*p*_*m*_; *Eff, Av*_*n*_) of dying at the peak of the impact of the complications is considered to be dependent on the natural probability inherent in the type of complication *p*_*m*_, i.e. on the severity of the illness triggered by the virus, and on the efficiency *Eff* and availability *Av*_*n*_ of medical help (see below).

Finally, in addition to *P*_0_(*k*) describing the probability that the virus is transmitted to a susceptible during the latent period or, if the infection carrier remains asymptomatic, during the time of his/her recovery, we need to introduce

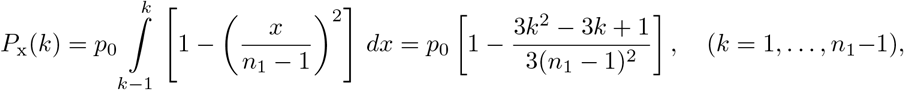

which is the probability of contracting the infection from a person with clinical symptoms. This is relevant to the medical staff who deal with such people, i.e. those who no longer take part in normal daily activities. To calculate this probability, we used the same algebraic form of the immune response as in (1).

### Vulnerability

In addition to categorizing the population with respect to their social role, we need to take into account that different categories of people may have different vulnerability to the disease in question. In order to account for this factor, we introduce a parameter *V*_*C*_, (*C* = *W*, …, *U*) considered to be in the range 1 ≤ *V* < ∞, with *V* = 1 corresponding to no specific vulnerability. For the categories introduced above, we have *V*_*W*_ = *V*_*S*_ = *V*_*M*_ = *V*_*U*_ = 1 and, generally, *V*_*R*_ *>* 1. Then, we will modify probabilities *P*_*j*_(*k*) (*j* = 0, …, 3) and instead use

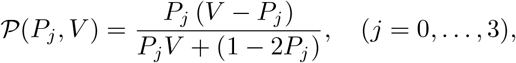

where the functional form of *þ* is chosen such that *þ* (*P*_*j*_, 1) = *P*_*j*_ and for *P*_*j*_ ≪ 1 the descriptive phrase ‘*n* times more vulnerable’ corresponds to *þ* (*P*_*j*_, *n*) ≈ *nP*_*j*_ and the emotional ‘infinitely more vulnerable’ corresponds to *þ* (*P*_*j*_,) = 1.

### Healthcare system

In terms of the present approach, the healthcare system impacts on the epidemic dynamics in two ways. The first is implicit, via the testing programme covering large sections of the population. It results in an increase of *p*_1_ due to detection and subsequent isolation of asymptomatic infectives and, as a consequence, in a reduction of *p*_2_ as the asymptomatic infectives ‘artificially’ brought from 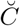 into the 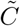 category (*C* = *W*, …, *U*) will not develop complications to become critically ill. This dual role of the testing programme can be either accounted for from the start by setting the appropriate values of the two probabilities or considered as a controlling factor in handling the epidemic.

The main role of the healthcare system is to reduce the death toll, i.e. reduce *p*_3_(*p*_*m*_, *Eff, Av*_*n*_). Besides its direct societal impact, this function of the healthcare system brings back into the community those who are now immune to the virus and hence contribute to creating the cushion distancing infectives from susceptibles thus suffocating the epidemic via the mechanism of herd immunity.

The probability *p*_3_ = *p*_3_(*p*_*m*_, *Eff, Av*_*n*_) depends on the natural mortality *p*_*m*_ and the efficiency *Eff* and availability *Av*_*n*_ of treatment. We will use a simple bilinear dependence

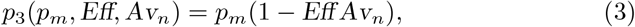

so that the fully available (*Av*_*n*_ = 1) and 100%-efficient (*Eff* = 1) medical help results in *p*_3_ = 0 whilst, if either *Eff* = 0 (no efficient treatment) and/or *Av*_*n*_ = 0 (medical help is unavailable), *p*_3_ = *p*_*m*_, as should be expected.

The value of *Eff* is determined by the current level of medical knowledge of the particular complications caused by the virus and the available treatment; it increases with the arrival of new more efficient ways of treating them and/or newly discovered applications of existing medicines. In the present approach, *Eff* is considered as an external control parameter.

By contrast, the availability parameter, *Av*_*n*_, depends on the current number of critically ill patients 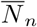, defined as

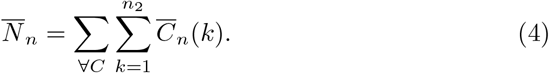

and the number of hospital beds, *B*_*n*_, available for them:

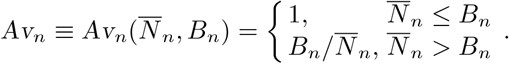

The term ‘hospital bed’ and the number of beds can mean different things for different illnesses. For example, for severe pneumonia the number of ‘beds’ can mean the number of hospital places in intensive care units fully equipped with ventilators, oxygenizers, etc and adequately staffed with qualified personnel. Thus, the number of ‘beds’ *B*_*n*_ depends on what we will collectively call ‘equipment’ *B*_eq_ and the patient-to-staff ratio 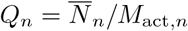, where *M*_act,*n*_ is the number of active medical staff defined as

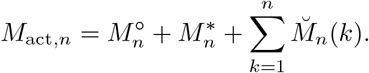

For *B*_*n*_ we will use the following dependence

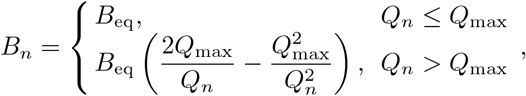

where *Q*_max_ is the maximum patient-to-staff ratio corresponding to normal (as opposed to ‘overstretched’) functioning of the healthcare system.

Another aspect of the healthcare system in the case of a viral infection is protection of the frontline medical staff who at work are most exposed to the infection. We will accumulate all protective measures in a parameter *N*_ppe_ (‘PPE’ stands for ‘personal protection equipment’), representing the ‘effective’ number of kits ensuring complete protection. Then the coefficient *X*_*n*_ characterizing the exposure of the medical staff to the hospital environment can be introduced as follows

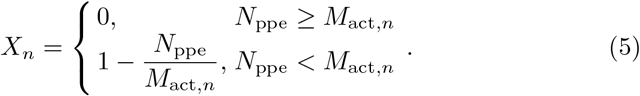

Together with *Eff*, parameters *N*_ppe_ and *Q*_max_ are external to the model and play the role of controlling factors.

It should be noted that the description of the healthcare system (above and below) can become important for relatively small communities whilst for countries mortality rarely becomes a factor influencing the dynamics of an epidemic so that one can use various simplifying assumptions to avoid going into detail of how the healthcare system is equipped.

## 4 Dynamics of epidemic

### 4.1 Transmission process

The conceptual framework outlined above largely followed the compartmental approach, albeit with considerably diversified compartments, and now we will bring in the stochastic element applying it to the compartments rather than individuals and linking it to the activities that individuals belonging to different categories go through on a daily basis.

The key idea in the modelling of the spreading phase is to consider every day of an epidemic as a succession of activities and evaluate the probability for a representative individual from category *C* (*C* = *W*, …, *U*) to pass through all these daily activities without being infected and hence ‘survive the day’. The activities are regarded as independent so that the probability of surviving the day is the product of probabilities of surviving each of the daily activities. Those who fail to survive the day become infected.

In other words, for every category *C* of the population the number of new cases ‘tomorrow’, i.e. on day (*n* + 1) of the epidemic, is given by

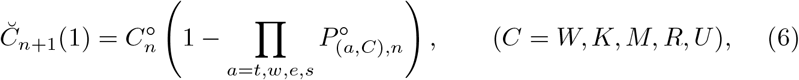

where 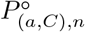 is the probability that ‘today’, i.e. on day *n*, a representative from *C* will survive activity *a*.

### Commuting by public transport

In this activity, the probability 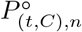, equal to the product of the probabilities of surviving all casual contacts in public transport, is the same for all categories:

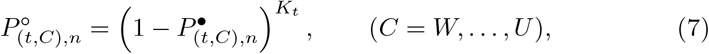

where 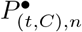 is the mathematical expectation of the probability of becoming infected as a result of one contact. For the latter, we have

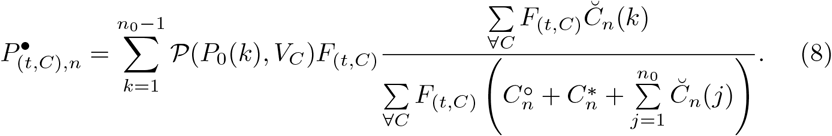

Here the last factor is the ratio of the number of the commuters who contracted the virus *k* days ago to the total number of people commuting by public transport, so that the fraction represents the probability of coming in contact with an infective should one use public transport. A similar expression albeit in a different context can be found e.g. in [19]. The preceding factor is essentially the probability of someone from *C* him/herself being on public transport, so that the product of the last two factors is the probability of an encounter between a susceptible from *C* and an infective. The first factor on the right-hand side of (8) is the probability of contracting the virus during such an encounter.

### Work

The categories we use here are defined with respect to one’s role in maintaining the living environment, so that we immediately have

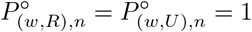

since *R* and *U* are economically inactive. For *W* we have (7) but with a different number of contacts,

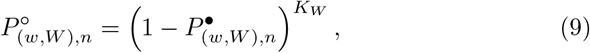

and for 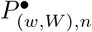 the expression is similar to (8) but involves a different set of people,

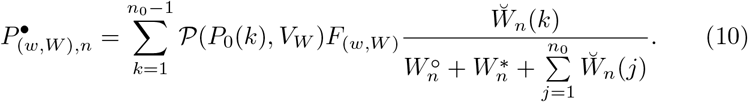

For category *S*, there are two types of interactions, with colleagues and with customers, so that, instead of (9), we now have

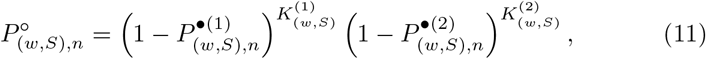

where, similarly to (10),

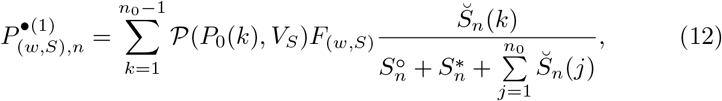

and, similarly to (8),

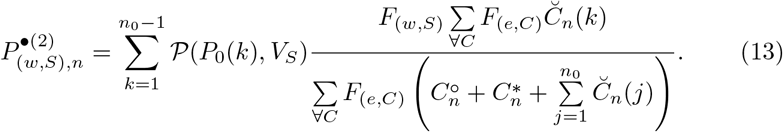

Finally, for category *M*, i.e. medical staff, who at work deal almost exclusively with infectives and hence have to rely on their personal protection equipment limiting their exposure to the virus, we have

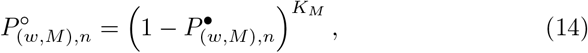

where

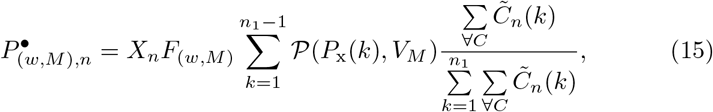

and, obviously, if there are no infectious patients yet, the 0*/*0 uncertainty on the right-hand side of (15) should be resolved by setting 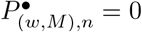.

### Interactions with services

All categories of the population also act as customers, so that, in a similar way as before, we have

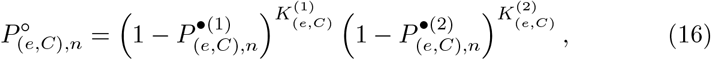

where

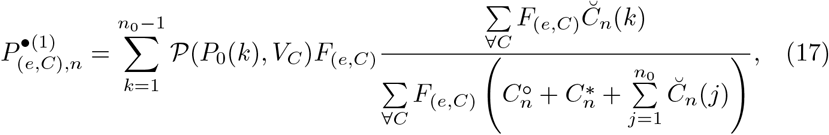

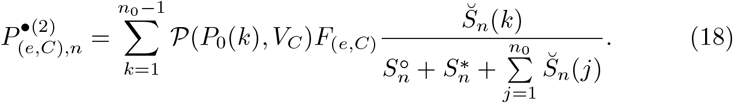

### Socializing

Socializing is as diverse as life itself, and here, with the broad categories we introduced, it will be represented schematically as socializing within one’s category, thus accounting for the fact that the bulk of after-work interactions often happen within one’s professional/social group, and, summarily, with all other categories taken together. This scheme of socializing makes this activity somewhat similar to the work pattern of the service providers considered above with the difference that the bulk of contacts happens with ‘colleagues’ from the same social group and to a less extent with ‘customers’ from the outside. Then, we have that for every category *C* (*C* = *W*, …, *U*)

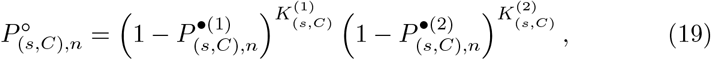

where

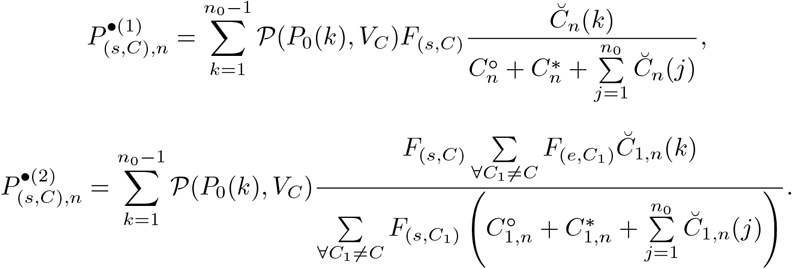

As a consequence of the transmission (6), we obviously have that ‘tomorrow’, i.e. on day (*n* + 1), there will be fewer susceptibles:

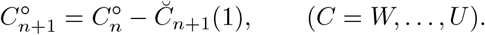

4.2 Illness, recovery and mortality

The number of those who become ill and hence withdraw from the daily activities is given by

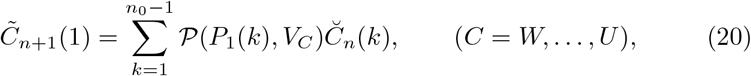

which accounts for the possibility of people showing clinical symptoms of the disease at different times within the incubation period. A direct consequence of (20) is the corresponding reduction in the number of asymptomatic infectives who continue their daily activities and hence continue infecting susceptibles:

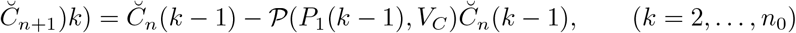

for *C* = *W*, …, *U*.

Transition from showing symptoms to becoming critically ill follows the same pattern as transition from being an asymptomatic carrier of the virus to becoming ill/symptomatic:

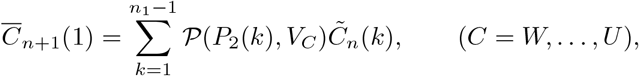

and hence

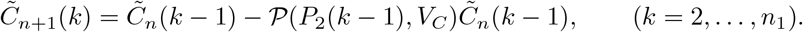

Finally, for the numbers of remaining critically ill, dying and recovering we have

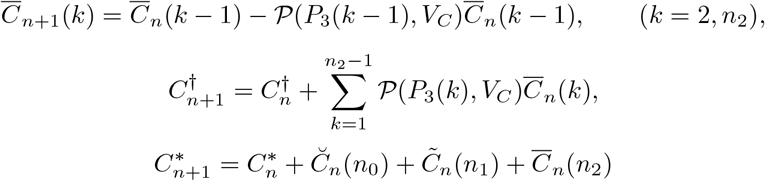

for *C* = *W*, …, *U*. The above algorithm involves probabilities, which are fractional, and the numbers in different categories, which are, of course, integers. Therefore, in the numerical implementation of the algorithm one should use the corresponding description of the variables so that fractions in the number of people are rounded to the nearest integer. One consequence of this obvious element is that, unlike the dynamical systems approach with continuous variation of the number of people in compartments, there is a clear end to an epidemic with no fractional lingering that needs an artificial ‘cut-off’. As for the number of contacts featuring as parameters in the present model, they should be understood as average and are, in general, fractional.

### Values of parameters

Unlike the dynamical systems type models, the approach outlined above requires no such concepts as ‘mixing matrices’ or ‘coefficient of variation’, whose values are difficult to assess not to mention prescribe. All parameters used here have clear meanings, and should fitting the model to the data require, say, the average number of contacts in the hundreds, this would immediately flag up the model’s inadequacy.

The map of social interactions involves two groups of parameters, the first are assembled in the participation matrix *F*_(*a,C*)_, where the entries vary between 0 and 1, whilst the second characterizes different types of contacts. The latter could be taken from surveys and at the time being plausibly assumed within a realistic range. Note that parameters from these two groups appear in different ways in the transmission process, the first as factors whilst the second as powers, which is indicative of the combination of the compartmental and stochastic approaches.

Parameters of the viral infection and the complications it causes also fall into two groups, the first comprising different time scales and the second one including probabilities. The conceptual framework we use allows one to map the clinical picture of a disease directly onto the set of the time scales. It should be noted only that in translating the statistical data one has to take into account that they are modified by the medical intervention so that, for example, for non-survivors the typical interval between showing clinical symptoms of the virus and dying of complications reported by statistics can be a bit longer than the ‘natural’ time *t*_*m*_ used in the model.

Of the set of probabilities, the most important is *p*_0_. On the semilogarithmic axes, as we will see below, the plots describing the numbers of the symptomatic, hospitalized/critically ill and the dead as functions of time all shadow the plot of the infectives (see below), and the slope of the linear section of these plots (see Fig. 1, right), for a given socializing pattern, is determined by *p*_0_. Hence the value of this probability can be determined from the initial semilogarithmic slope of the statistical data reported at the beginning of an epidemic and then used to predict its course.

The application of the model to a specific viral disease is described below.

## 5 Illustrative example: COVID-19

We will illustrate how the simplest model developed in the framework of the new approach works using the ongoing pandemic of the coronavirus disease (COVID-19) as a convenient example and the UK as a representative country. On the one hand, although generating an avalanche of papers this disease is yet not as well studied as some past epidemics and even the timeline in its clinical features reviewed in a number of publications, e.g. [26, 32, 33], is still not entirely settled. On the other hand, however, the global nature of this pandemic triggered a variety of non-pharmaceutical measures (social distancing, quarantines, etc) aimed at curbing the outbreak, some unprecedented in their scale, and this makes it possible to test, at least on a qualitative level, the model where these measures can be accounted for in a straightforward way by comparing its predictions with this ongoing ‘experiment’.

We will try to fit theoretical curves to the data reported in the UK for both the number of cases, interpreted this number, according to the guidelines issued by the UK Governmane, as the number of patients 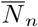 defined by (4)), and the total number of deaths. It might seem that the number of parameters available in the model should allow one to fit almost anything but this is not the case. After we incorporate all what is known about the disease, thus eliminating all degrees of freedom associated with the time scales involved, and use natural restrictions on the numbers specifying social contacts and their variation due to the imposed lockdown, almost all flexibility in the model is gone, so that essentially it is only some probability scales that remain and even their range is roughly known from the clinical data. The procedure is described below to make our calculations reproducible.

### Pre- and post-lockdown social signatures

According to the Office for the National Statistics (UK), out of the population of about 66.65m we have *R* = 12m, *U* = 9.9m whilst the remaining population, including 4.5m of health and social care workers, has to be ascribed to categories *W, S* and *M* using the appropriate assumptions and guidance from the ONS breakdown into industries. Of all service industries, we will attribute to category *S* retail, transport, food, financial services,real estate social care and some other services. The estimated total is *S* = 14.5m. For the frontline medical staff we take *M* = 1.2m. The remainder is attribute to *W*, i.e. those whose work or education involve interacting within their community: *W* = 29.05.

For the broad categories we use here, in the absence of the relevant sociological data regarding interaction patterns of different social groups, the participation matrix *F*_(*a,C*)_ can only be assumed on the basis of general considerations. For the pre-lockdown period, we will assume

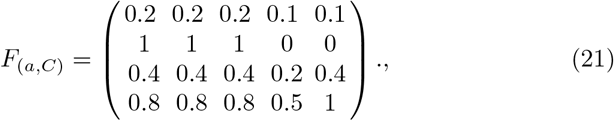

where the entries for *a* = *t, e* come from estimates for the time spent on average in the corresponding activity; for *a* = *s* for the main working groups (*C* = *W, S, M*) they are taken as comparable with work given that we do not consider a separate participation matrix for weekends. For the contacts we set: *K*_*t*_ = 4, *K*_*W*_ = 5, *K*_*M*_ = 10, 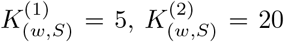 and 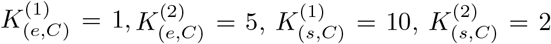 for *C* = *W*, …, *U*. The value of *K*_*M*_ is actually irrelevant as for simplicity we will assume that medical staff have an adequate supply of personal protection equipment (*N*_ppe_ ≥ *M*), so that their exposure to the virus at work *X*_*n*_ = 0.

On Monday 23 March 2020, a lockdown has been announced and nonessential business activities, including closures of much of retail, schools, universities, pubs, hairdresses, etc, have been closed. This was accompanied by measures of social distancing imposed on outdoors socializing. We will assume that within the week 23–30 March 2020 the normal participation matrix (22) linearly evolved into the following one

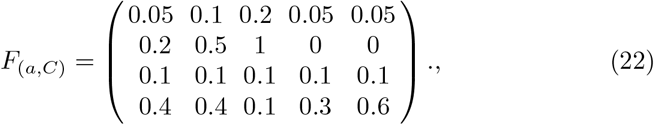

where the reduction in work for category *W* and to a less extent *S* is accompanied by the corresponding reduction in the use of public transport; interaction with services is restricted to the essentials and socializing is halved. As for the number of contacts, the only assumption we make is that casual interactions with fellow shopper is now absent: 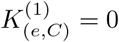, *C* = *W*, …, *U*.

### Time scales associated with the virus

According to [34], the incubation period of COVID-19 is 3 days for high viral load and 4 days otherwise, whilst other works reported that it is 5 days [32, 35], and in extreme cases can be between 1 and 7 days [35] or even longer [32]. For our computations, we set *t*_1_ = 4, *σ*_1_ = 1.5, which is consistent with the statistical analysis of the publicly available data reported in [36].

The value of *n*_0_, which describes the duration of the illness in asymptomatic carriers, is somewhat difficult to discern. However, for COVID-19 we have the results of a thorough field study [37] conducted by a joint Italian-British team in the municipality of Vo’, Italy, where an entire community of nearly 3,000 residents was locked down and monitored for two weeks. The study found a surprisingly large (43.2%) proportion of all cases being asymptomatic and reported ‘the time of viral clearance’ and the ‘positivity window’ to be 9.3 and 9.1 days, respectively. On the basis of these findings, in our computations we set *n*_0_ = 10.

The time scale of recovery for symptomatic patients from the viral infection influences only the return of now immune individuals back into the community, i.e. plays a minor role in the overall dynamics of the epidemic when the number of infectives is much larger than of those recovered, so for *n*_1_ we simply take the value from the chart collated in [33] and set *n*_1_ = 13.

Clinically, in the adverse case, fever and cough are followed by dyspnea, ARDS and death. The median duration from the first symptoms to dyspnea is 5 days [25] and to death is 10 days [38] with artificial ventilation and other measures capable of prolonging it to 14 days [39]. Thus, in our computations we set *t*_2_ = 5, *t*_*m*_ = 5.

### Interpretation of reported data

As already mentioned in Section 2, the UK guidance to potential COVID-19 sufferers prescribed that they should neither go to their GP nor to hospital; instead they should stay indoors, get advice online and call the ambulance if their condition gets worse or the symptoms do not get better after seven days. So, in terms of our model, the reported cases should be interpreted as those who did not recover on their own and ended up in hospital, i.e. joined 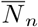 defined by (4). Thus, we will interpret the reported cases as 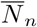.

### Scales for probability distributions

The scales for the probability distributions can be related to the post factum integral probabilities reported in the epidemic statistics only via the application of the model itself, so, subject to some restrictions, we will be using these scales as adjustable parameters in the following way.

With the social signature of the population (*F*_*a,C*_ and the contacts) fixed, the probability *p*_0_ determines the initial exponential growth of the number of cases, i.e. the slope of the linear region in the semilogarithmic plot (Fig. 1, right). Given that *p*_1_ determines, roughly, the percentage of asymptomatic cases, to make the outcome consistent with the reported data [37], we set *p*_1_ = 0.65. The scales *p*_2_ and *p*_*m*_ determine the probability of the virus causing complications and the probability of dying should there be no medical help, respectively. We will assume *p*_2_ = 0.046, consistent with reported estimates [26] but ultimately an adjustable value here. As for *p*_*m*_, it is mitigated by the efficiency and availability of medical help, i.e. *Eff* and *Av*_*n*_ (3), and we will assume that medical help is available without restriction, *Av*_*n*_ = 1, and split *p*_*m*_ and *Eff* as follows: *p*_*m*_ = 0.2 and *Eff* = 0.5, which makes the actual probability of dying *p*_3_ = 0.1 consistent with the top of the 4–11% range reported in [26] for hospitalized adult patients.

We need also to take into account that elderly people forming our category *R* are more vulnerable than other categories, according to [38, 26], approximately three times, so we set *V*_*R*_ = 3 whilst *V*_*W*_ = *V*_*S*_ = *V*_*M*_ = *V*_*U*_ = 1.

For our computations, we introduce a small number 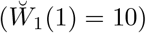 of infectives and, to fit the data starting from the beginning of March 2020, shift the time by 35 days, which, incidentally, takes us right into the middle of the Milan winter sales, where, as reported, COVID-19 has been brought by tourists from China.

### Reported data, retrospection and prediction

Fig. 3 shows the cases and deaths reported in the UK versus the corresponding curves computed theoretically with the assumptions listed above. For *Eff* = 0.5 for the whole period, the theoretically computed death toll is slightly higher than reported but if we factor in an improvement of health care as experience is gained and use *Eff* = 0.7 from the beginning of May 2020, the match becomes perfect.

**Fig. 3.**
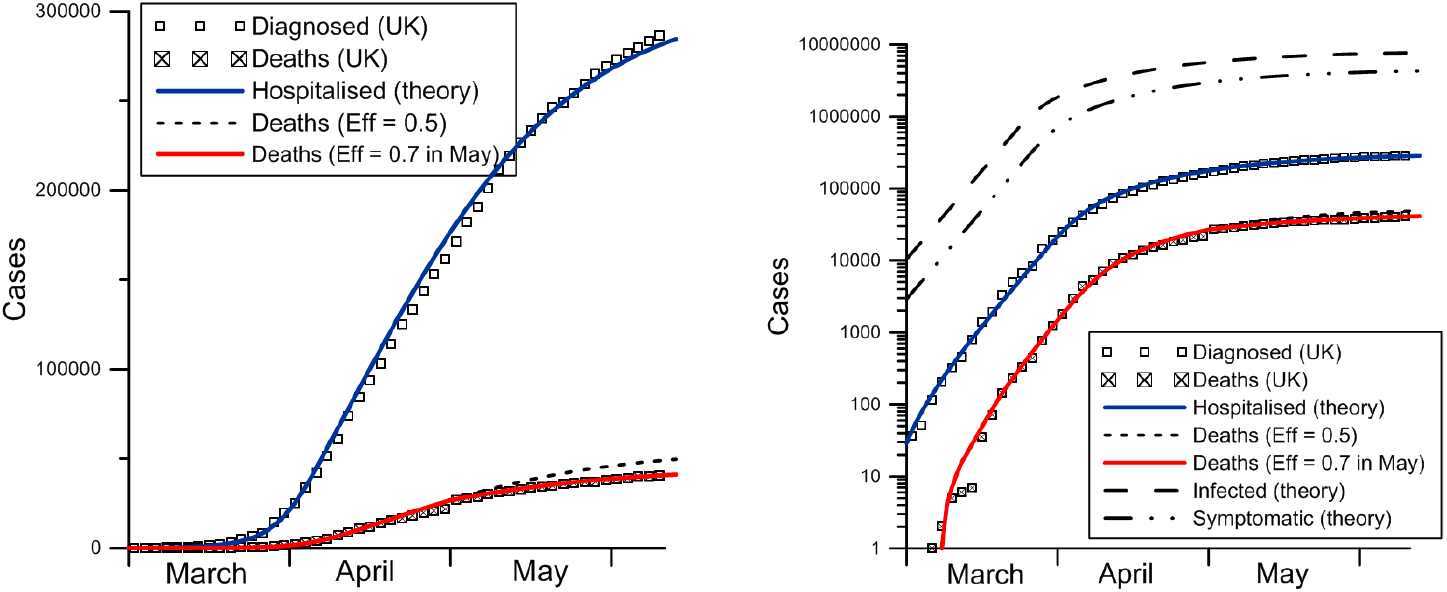
Diagnosed cases and fatalities reported for COVID-19 in the UK in the spring of 2020 and theoretical curves on the linear (left) and semilogarithmic (right) axes. The two variants of the theoretical death toll correspond to *Eff* = 0.5 for the whole period and for *Eff* = 0.7 from 1 May 2020. The right figure shows also theoretically obtained numbers of infectives and symptomatic/mild cases. The parts of the curves corresponding to February are not shown.

As one can see, on semilorarithmic axes (Fig. 3, right), the curves showing the evolution of the symptomatic, hospitalized and fatal cases broadly shadow the curve of the infectives. It is noteworthy that the model allows one to describe both the initial stage (more pronounced on the semilogarithmic plot) and the later stage (more visible on the linear plot). Obviously, our comparison of the theory with observational data is nothing but a successful attempt of curve-fitting, albeit with all information about the disease and plausible assumption used as restrictions. What we can do now is to fix all the values that allowed us to describe the data and apply the theory, first, for a retrospective analysis.

Consider retrospectively two scenaria alternative to what actually happenned, i.e., the lockdown imposed on 23 March 2020 The first scenario is ‘do nothing’, i.e. the Swedish variant with no lockdown at all, and the second one is lockdown imposed a week earlier, i.e. from 16 March instead of 23 March 2020. In both scenaria, we will use *Eff* = 0.7 from the beginning of May which, as we have already seen, gives a more accurate description of the actual death toll. The fatalities in these two scenaria are shown in Fig. 4, again as linear and semilogarithmic plots. One can see that the do-nothing strategy would have led to the death toll more than seven times higher than the actual one. At the same time, should the lockdown be imposed a week earlier, the number of deaths would have been reduced by about 60%.

**Fig. 4.**
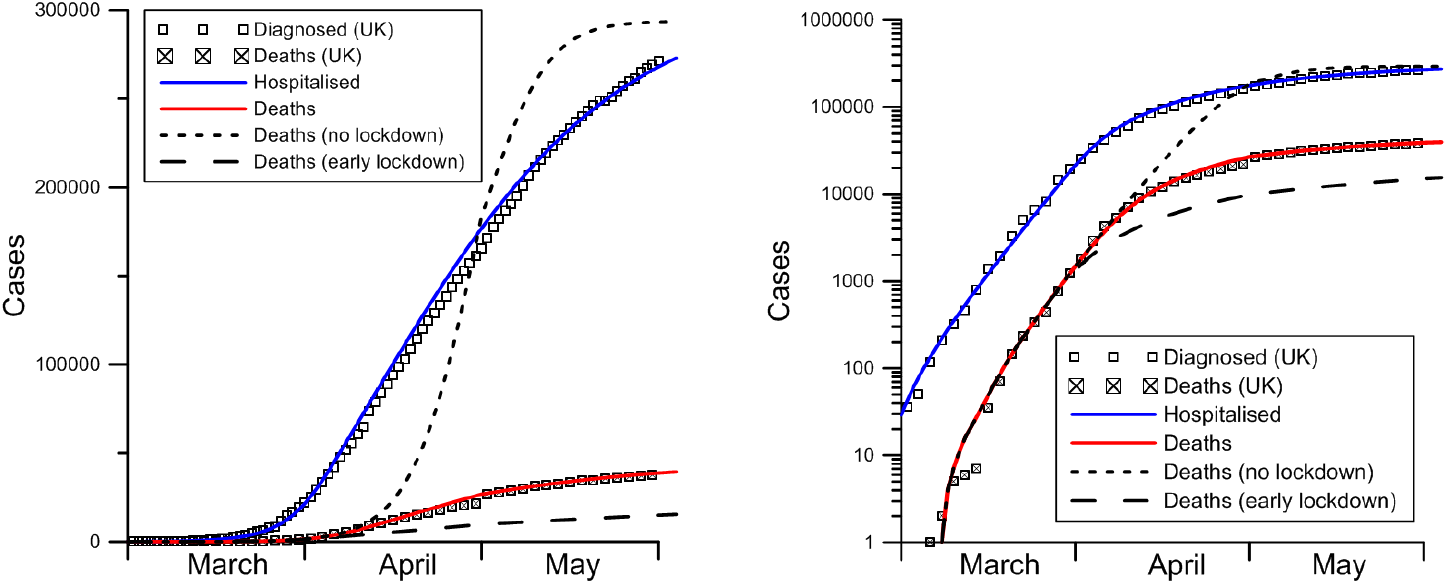
The fatalities in the do-nothing (no lockdown) and an earlier lockdown scenaria are added to the data and theoretical curves of Fig. 3.

A qualitative conclusion one can draw from these findings is that at the beginning of an epidemic, when the number of cases grows exponentially, the time lag between contraction of the virus and its consequences in terms of complications and deaths plays a crucial role, and, with a large percentage of asymptomatic and (unreported) mild cases, it is important to curb this exponential growth early as the diagnosed/severe cases are a belated indicator of the state of the epidemic.

It is worth noting also that the timing of the peak daily mortality rate in the do-nothing scenario was predicted in March 2020 in a report [40] by the Imperial College COVID-19 Response Team on the basis of the then available information and an earlier developed model [41]. According to their model, which focusses on the population location distribution aspect, the peak was expected at the beginning of June 2020, whilst, as one can see in Fig. 4 (left), the present model, which operates with social and economic activities rather than locations, places the peak in late April 2020. The total death toll for the UK in the do-nothing scenario predicted in [40] was 510,000 whilst the present model puts it below 300,000. Both predictions are, of course, unverifiable.

The lockdown obviously cannot last forever, and it is interesting to look at what one should expect if it is lifted altogether, with the population returning (gradually, e.g. over a week) to its normal pattern of behaviour. This is shown in Fig. 5 for two variants: (i) the lockdown lifted on 1 June and (ii) on 15 June 2020.

**Fig. 5.**
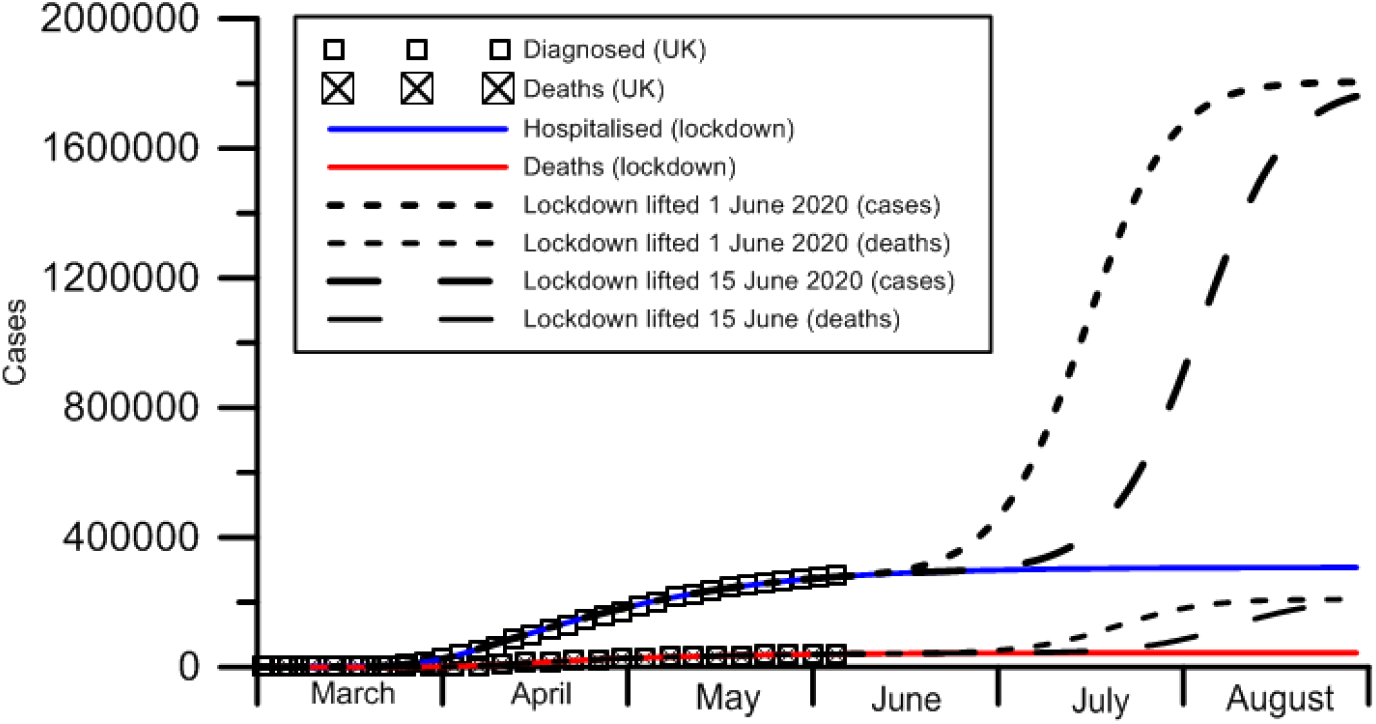
The number of cases and deaths for the scenaria of the lockdown lifted on 1 June and on 15 June 2020.

As one can see from this figure, there is a potential for a second wave of the epidemic, with a peak approximately 1.5 months after the lockdown is lifted altogether. Notably, the total death toll corresponding to each variant of the lockdown lifting is considerably lower than what one would have had should the do-nothing strategy be adopted from the start. In other words, social distancing and quarantine measures, temporary as they are, help to reach herd immunity with considerably lower number of fatalities.

It should be also noted that COVID-19 mutates [42] and, according to some studies [34], has started to become milder even within China, which is another argument in favour of social distancing and other non-pharmaceutical protective measures used to curb the epidemic. The effect of the weather is found to be inconclusive [43] which makes hopes that, regardless of any actions, the epidemic will simply go away unfounded.

The variation in the virus aggressiveness due to its mutations and other factors can be easily factored in the model via the scales for the probabilities it uses but we will not do this here.

## 6 Discussion

### 6.1 The modelling approach

The approach formulated above makes it possible to develop mathematical models of increasing complexity in a regular way, essentially by converting the data from sociological surveys of behavioural patterns of different groups of the population into the map of social interactions (the participation matrix and the numbers of contacts), thus describing the spreading of epidemics in more and more detail. The basis of the approach is that it essentially follows one’s daily routine (averaged over a suitable short period, say, a week), looking at it through the prism of one’s communications with different groups of people that might result in contraction of a communicable infection. The broad categories used in the simplest model we introduced here to illustrate the approach can be split into subcategories, if the available information allows, down to particular businesses, and account for the gender, age group, etc. Similarly, the set of activities can be made more detailed, most importantly regarding the socializing element where social contacts enable the disease to cross boundaries between categories. Basically, our approach follows the tracking method used to trace the interaction path of an individual infection carrier at an early stage of an epidemic. This tracking is the key idea of the stochastic approach, and here we applied it to larger cohorts of people, thus utilizing the compartmental framework. In this sense, the new approach unifies the two main approaches that so far have been developing separately.

The rationale behind the chosen way of categorizing the population is that, once an epidemic is declared and the authorities are forced to take action, the shutdown of businesses and various quarantine measures will hit the economy depending on how it is structured in terms of its functioning and in terms of the corresponding communication pattern. This feature is the key difference between the new approach and the models treating the population as structureless, essentially animal-like, solely in terms of compartments related to the disease (SIR, SEIR, etc). An immediate consequence of the difference is that the recommendations following from the latter apply indiscriminately to the whole population irrespective of what their economic implications may be thus essentially making them unworkable. By contrast, the present framework makes it possible to rank the categories it uses with regard to their importance for the short- and medium-term functioning of the economy and thus allows one to produce a ‘map’ of an epidemic with the two axes being the cost to society due to the disease and the cost due to shutting down the economy. This map, broadly similar for different communicable diseases, would help to strike the socially acceptable balance between these two costs.

The natural next step in developing the simplest model formulated in the framework of the new approach is to bring in the notion of ‘household’. From the viewpoint of our approach, the household is simply a place where intensive socializing between representatives, potentially, of different categories takes place. In other words, this notion can be incorporated without any ad hoc constructions. However, for the resulting model to be specific and hence useable, one needs sociological information about the ‘profile’ of households for a given population in terms of representatives of what categories and in what proportion households bring together, as it is this information that largely determines the probabilities of transmission of the disease between different categories.

There are two elements in the new approach that remain to be developed conceptually. The first one is the geographic aspect. For a vast country, like Russia or the US, the population mixes globally on a larger time scale than that of an epidemic, so that one should use the models similar to the simplest illustrative model described here for intensively mixing local communities (cities, regions, or, in the case of the US, states) and introduce flows of people between them. Such flows can be introduced in an obvious way but, for the embracing model to be specific, one will need information regarding how travelling between different locations is inscribed in the daily/weekly routines of the people belonging to different categories.

The second element to be incorporated is the social activity of ‘super- spreaders’. Superspreaders form a subcategory of people hyperactive in their social contacts. Observational data on COVID-19 show that one superspreader can create a cluster of tens of infectives or even, going on a socializing spree, infect more than a hundred people across different categories in a matter of days. The development of our approach in this direction is technically more challenging as it requires profiling of the population with regards to their social activities and the corresponding modification of both the participation matrix and the set of contacts.

### 6.2 The simplest model and COVID-19

As shown, the simplest model formulated in the framework of the new approach can be fitted to the observational data of the COVID-19 epidemic in the UK, simultaneously for both the number of cases and the number of deaths both before and after the lockdown, with somewhat surprising accuracy. Furthermore, in this fitting, much of the flexibility in the model’s parameters has been removed by incorporating the experimental data characterizing the virus as well as by additional constraints following from the meaning of the parameters accumulated in the map of socializing.

Although this comparison with the data as well as the simplest model itself were intended only as an illustration of the new modelling approach, this comparison and computations run using the model allow one to draw some qualitative and, to some extent, quantitative conclusions regarding the epidemic. The first one is that shutdown of non-essential businesses, social distancing and other non-pharmaceutical measures taken to curb the epidemic not only slowed down the spread of the disease but also, as shown, reduced the herd immunity level, i.e. the proportion of symptomatic cases in the population needed for the epidemic to come to an end. For COVID-19, this level is found to be considerably lower than what is normally expected for viral infections. This result follows from the conclusions of the field study [37] that a large proportion of asymptomatic cases plays a key role in the spreading of COVID-19.

It has also been shown that, should the non-pharmaceutical measures in the UK be imposed just a week earlier, the death toll by now would have been considerably lower, approximately by 60%. In other words, such measures should be imposed at the beginning of the exponential phase of an epidemic as their effect later on becomes greatly diminished.

Finally, for COVID-19, the model shows that there is a potential for the second wave of the epidemic, and the magnitude of this wave decreases the longer the measures of social distancing remain in place.

## Data Availability

All data used in the work are publicly available.

## Acknowledgements

The authors wish to thank Grant Armstrong for inspiration and helpful remarks and Penelope Barber for resolving some technical issues that COVID-19 brought about.

## References

1 F. Brauer, Infectious Disease Modelling 2, 113 (2017)

2 W.O. Kermack, A.G. McKendrick, Proceedings of the Royal Society of London A: Mathematical, Physical and Engineering Scinces 115, 700 (1927)

3 W.O. Kermack, A.G. McKendrick, Proceedings of the Royal Society of London 138, 55 (1932)

4 W.O. Kermack, A.G. McKendrick, Proceedings of the Royal Society of London 141, 94 (1933)

5 H.W. Watson, F. Galton, J. Anthrop. Inst. Great Britain and Ireland 4, 138 (1874)

6 F. Galton, Natural Inheritance (Macmillan, 1889)

7 G.F. Steffensen, Matematisk Tiddskrift B 1, 19 (1930)

8 G.F. Steffensen, Annales de l’institut Henri Poincaré 3, 319 (1931)

9 J.A.J. Metz, Acta Biotheoretica 78, 75 (1978)

10 O. Diekmann, J.A.P. Heesterbeek, Mathematical Epidemiology of Infectious Diseases: Model Building, Analysis and Interpretation (John Wiley and Sons, Inc., 2000)

11 M. Choisy, J.F. Guégan, P. Rohani, in Encyclopedia of Infectious Diseases: Modern Methodologies, ed. by M. Tibayrenc (John Wiley and Sons, Inc, Hoboken, New Jersey, USA, 2007), pp. 379–404

12 A. Lajmanovich, J. Yorke, Math. Biosci. 28, 221 (1976)

13 L. Sattenspiel, C.P. Simon, Math. Biosci. 90, 341 (1988)

14 J. Dushoff, L. S., Math. Biosci. 128, 25 (1995)

15 J. Arino, in Modeling and dynamics of infectious diseases, ed. by Z. Ma, Y. Zhou W. J. (World Scientific Publ., Singapore, 2009), pp. 65–123

16 B. Bonzi, A. Fall, A. Iggidr, G. Sallet, J. Math. Biol. 62(1), 39 (2011)

17 S.P. Blythe, C. Castillo-Cavez, Math. Biosci. 96, 221 (1989)

18 J.A. Jacqucz, C.P. Simon, J. Koopman, in Epdemics models, their structure and relation to data, ed. by D. Mollison (Cambridge University Press, Cambridge, 1996), pp. 279–301

19 D. Bichara, A. Iggidr, J. Math. Biol. 77, 107 (2018)

20 D. Bichara, Y. Kang, C. Castillo-Chavez, R. Horan, C. Perrings, Bull. Math. Biol. 77, 2004 (2015)

21 D. Bichara, S.A. Holechek, J. Velázquez-Castro, A.I. Murillo, C. Castillo-Chavez, Lett. Biomath. 3, 140 (2016)

22 D. Bichara, C. Castillo-Chavez, Math. Biosci. 281, 128 (2016)

23 C. Castillo-Chavez, D. Bichara, B.R. Morin, Proc. Natl Acad. Sci. 113, 14582 (2016)

24 G.K. Batchelor, An Introduction to Fluid Dynamics (Cambridge Univ. Press, 1967)

25 W. Wang, J. Tang, F. Wei, J. Med. Virol. (2020). DOI 10.1002/jmv.25689

26 T. Singhal, Ind. J. Pediatrics 87, 281 (2020)

27 I.C. Marschner, Math. Biosci. 109, 39 (1992)

28 M. Morris, Sociol. Methods & Research 22, 99 (1993)

29 A.L. Lloyd, R.M. May, J. Theor. Biol. 179, 1 (1996)

30 C.O. Uche, R.M. Anderson, IMA J. Math. Appl. in Medicine & Biol. 13, 23 (1996)

31 J. Cui, Y. Zhang, Z. Feng, J. Biol. Dyn. 13, 31 (2019)

32 F. He, Y. Deng, L. W., J. Med. Virol. (2020). DOI 10.1002/jmv.25766

33 P. Kakodkar, N. Kako, M.N. Baig, Cureus 12, e7560 (2020). DOI 10.7759/cureus.7560

34 X.W. Xu, X.X. Wu, X.G. Jiang, K.J. Xu, L.J. Ying, C.L. Ma, S.B. Li, H.Y. Wang, S. Zhang, H.N. Gao, J.F. Sheng, Y.Q. Cai, H.-L. Qiu, L.J. Li, British Med. J. 368 (2020). DOI 10.1136/bmj.m606

35 M. Zhou, X. Zhang, J. Qu, Front. Med. (2020). DOI 10.1007/s11684-020-0767-8

36 N.M. Linton, T. Kobayashi, Y. Yang, K. Hayashi, A.R. Akhmetzhanov, S.m. Jung, B. Yuan, R. Kinoshita, H. Nishiura, MedRxiv pp. 1–11 (2020). DOI 10.1101/2020.01.26.20018754

37 L. Lavezzo, E. Franchin, C. Ciavarella, G. Cuomo-Dannenburg, L. Barzon, C. Dell Vecchio, L. Rossi, R. Manganelli, A. Lorengian, N. Navarin, D. Abate, M. Sciro, S. Merigliano, E. Decanale, M.C. Vanuzzo, F. Saluzzo, F. Onelia, M. Pacenti, S. Parisi, G. Carretta, D. Donato, L. Flor, S. Cocchio, G. Masi, A. Sperduti, L. Cattarino, R. Salvador, K.A.M. Gaythorpe, Imperial College London COVID-19 Response Team, A.R. Brazzale, S. Toppo, M. Trevisan, V. Baldo, C.A. Donnelly, N.M. Ferguson, I. Dorigatti, A. Crissanti, MedRxiv (2020). DOI 10.1101/2020.04.17.20053157

38 Korean Society of Infectious Diseases, Korea Centers for Disease Control and Prevention, J. Korean Med. Sci. (2020). DOI 10.3346/jkms.2020.35.e132

39 P. Sun, X. Lu, C. Xu, W. Sun, B. Pan, J. Med. Virol. 92, 548 (2020). DOI 10.1002/jmv.25722

40 N.M. Ferguson, D. Laydon, G. Nedjati-Gilani, N. Imai, K. Ainslie, M. Baguelin, S. Bhatia, A. Boonyasiri, Z. Cucunubá, G. Cuomo-Dannenburg, A. Dighe, I. Dorigatti, H. Fu, K. Gaythorpe, W. Green, A. Hamlet, W. Hinsley, L.C. Okell, S. van Elsland, H. Thompson, R. Verity, E. Volz, H. Wang, Y. Wang, P.G.T. Walker, C. Walters, P. Winskill, C. Whittaker, C.A. Donnelly, S. Riley, A.C. Ghani, Impact of non-pharmaceutical interventions (NPIs) to reduce COVID-19 mortality and healthcare demand. Tech. rep. (2020). DOI 10.25561/27482

41 N.M. Ferguson, D.A.T. Cummings, C. Fraser, J.C. Cajka, P.C. Cooley, D.S. Burke, Nature 442, 448 (2006). DOI 10.1038/nature04795

42 T. Phan, Infection, Genetics and Evolution p. 104260 (2020)

43 P. Byass, Global Health Action 13, 1760490 (2020)

